# Joint Associations of Plasma Nutritional Biomarkers and Uterine Fibroids with Hypertensive Disorders of Pregnancy

**DOI:** 10.64898/2026.05.12.26353013

**Authors:** Ananya S. Dewan, Mengmeng Li, Xiaobin Wang, Katherine Cameron

**Affiliations:** Department of Gynecology & Obstetrics, Johns Hopkins School of Medicine, Baltimore, MD 21205; Center on Early Life Origins of Disease, Department of Population, Family and Reproductive Health, Johns Hopkins Bloomberg School of Public Health, Baltimore, MD 21205; Department of Pediatrics, Johns Hopkins School of Medicine, Baltimore, MD 21205

## Abstract

**Background:** Hypertensive disorders of pregnancy contribute substantially to maternal morbidity and mortality, and occur with increased frequency among women with uterine fibroids. Biomarkers involved in oxidative stress and endothelial function, including folate, vitamin B12, vitamin D, and homocysteine, have been studied in relation to hypertensive disorders of pregnancy, but their relationship to fibroid-associated risk has not been well characterized, particularly in racially and ethnically diverse populations.

**Study Design:** This study was a retrospective analysis of the Boston Birth Cohort, a prospective cohort recruited at a large urban medical center. The analytic sample included 722 women with complete data on hypertensive disorder status, uterine fibroid status, and plasma biomarker measurements. Uterine fibroids and hypertensive disorders of pregnancy were ascertained through physician-assigned diagnostic codes and ultrasound report review. Plasma folate, vitamin B12, vitamin D, and homocysteine were measured in maternal or cord blood and analyzed as continuous variables and quartiles. Multivariable logistic regression models were used to estimate independent associations, evaluate interaction terms, and assess joint exposure categories.

**Results:** Of the 722 participants, 12% (86/722) had uterine fibroids and 10% (72/722) had a hypertensive disorder of pregnancy. Plasma micronutrient concentrations did not differ significantly by fibroid status. Women with hypertensive disorders of pregnancy had higher plasma homocysteine concentrations compared with those without (p=0.028). Hypertensive disorders of pregnancy were more common in the lowest folate quartile compared with the highest quartile (p=0.018) and in the highest homocysteine quartile compared with lower quartiles (p=0.031). In joint-effects analyses, higher odds of having a hypertensive disorder of pregnancy were observed among women with both uterine fibroids and low folate compared with women without fibroids and with adequate folate (p=0.027). No significant joint associations were observed for vitamin D, vitamin B12, or homocysteine.

**Conclusion:** In this cohort, the co-occurrence of uterine fibroids and lower folate concentrations was associated with hypertensive disorders of pregnancy. This joint exposure delineates a subgroup that may be clinically relevant for future studies aimed at refining maternal risk characterization and exploring targeted nutritional supplementation strategies.

## Introduction

Hypertensive disorders of pregnancy (HDPs) affect approximately one in seven delivery hospitalizations in the United States,^1^ and constitute a leading cause of maternal and fetal morbidity and mortality globally.^2^ HDPs encompass a spectrum of diseases, including chronic hypertension with superimposed preeclampsia (PE); PE; eclampsia; and hemolysis, elevated liver enzymes, and low platelets (HELLP) syndrome.^3^ In the United States, HDP-associated morbidity and mortality disproportionately affect Black women due to disparities in access to care, socioeconomic differences, and the higher prevalence of HDP risk factors.^1,4–6^

The pathophysiology of HDPs is likely multifactorial, including inadequate uteroplacental perfusion, immunologic changes, and endothelial dysfunction.^7,9^ Interestingly, benign pelvic tumors known as uterine fibroids (UFs) have been recognized to confer a two- to three-fold higher risk for HDPs.^10–12^ This association may reflect shared underlying mechanisms implicating inflammation, oxidative stress, and endothelial dysfunction. One possible shared contributing factor is 25(OH)D (vitamin D) deficiency, which has been associated with a greater risk of both UFs and HDPs.^13^

Beyond vitamin D, homocysteine, a circulating marker of oxidative stress, may represent a mechanistic link between plasma nutritional biomarkers and both UFs and HDPs. Folate and vitamin B12 serve as cofactors in homocysteine metabolism,^18^ and deficiencies in either nutrient can lead to elevated homocysteine concentrations and subsequent endothelial injury.^19^ While studies examining homocysteine and UFs have reported mixed or inverse associations, elevated homocysteine levels have been more consistently associated with an increased risk of HDPs, particularly preeclampsia.^20–25^ In contrast, no association has been observed between UFs and folate or vitamin B12,^14^ and findings regarding their associations with PE have been inconsistent.^15–17^

This study examined whether micronutrient status, a measurable, cost-effective, and clinically actionable factor, could modify the association between UFs and PE. Importantly, prior work investigating HDPs and micronutrient status has limited representation of racially diverse populations, limiting the generalizability of those results.^26–29^ To address this gap, this retrospective cohort study had two aims: 1) to assess the individual associations between plasma biomarkers (vitamin D, vitamin B12, folate, and homocysteine) with HDPs and UFs in a racially and ethnically diverse cohort; and 2) to determine if plasma biomarkers modify the association between UFs and HDPs.

## Methods

### Cohort

This study was a secondary analysis of the Boston Birth Cohort (BBC): a cohort prospectively recruited from 1998 with continuous enrollment at the Boston Medical Center, which serves predominantly urban, low-income, and racially and ethnically diverse patients.^30^ Previous studies have described the BBC and associated methods in-depth,^30–34^ which are briefly summarized below. The BBC initially enrolled women who delivered singleton live infants who were preterm (<37 weeks) or low birthweight (<2500g), with matched term (≥37 weeks) and normal birthweight (≥2500g) controls.^30^ Exclusion criteria included women with pregnancies conceived with in vitro fertilization, multiple gestations, chromosomal abnormalities, or major birth defects.^30^ Written informed consent was obtained from each mother. The BBC has been approved by the Institutional Review Boards of the Johns Hopkins Bloomberg School of Public Health, Baltimore, MD, and the Boston Medical Center, Boston, MA.

Over 90% of eligible women approached by the BBC team were enrolled, resulting in 8623 total participants.^31^ A subset of women with complete data for HDP status, UF status, cord vitamin D, plasma B12, and plasma folate (n=722) was included in this analysis.

### Measures

At delivery, cord blood samples were collected. Cord blood plasma concentrations of 25(OH)D2 and 25(OH)D3 were measured using high-performance liquid chromatography tandem mass spectrometry (HPLC-MS/MS) and summed to derive the total 25(OH)D concentration.^32^ In this study, cord blood total 25(OH)D concentration serves as a proxy of maternal vitamin D status.

Maternal blood samples were collected 24-72 hours after delivery.^33^ Plasma homocysteine levels were measured using automatic clinical analyzers (Beckman-Coulter), whereas plasma folate was analyzed via chemiluminescent immunoassay with diagnostic kits (Shenzhen New Industries Biomedical Engineering Co., Ltd., China).^33^ Plasma vitamin B12 was measured using the Beckman Coulter ACCESS Immunoassay System (Beckman-Coulter Canada, Mississauga, Canada) with a MAGLUMI 2000 Analyzer.^33^

HDP and UF status were abstracted from the maternal medical record based on International Classification of Diseases (ICD) 9 or 10 diagnoses recorded by physicians before or during pregnancy, and review of obstetric or gynecologic ultrasound reports one year before or during pregnancy.^12,35^ HDPs included chronic hypertension with superimposed PE, PE, eclampsia, and HELLP syndrome. Definitions for each condition can be found in prior publications on the BBC.^35^

Self-identified race and ethnicity, highest level of education, and smoking were gathered from a standardized maternal questionnaire interview.^30^ Self-reported pre-gestational weight (kg) was divided by (height (m))^2^ to calculate body mass index (BMI).^12^ Maternal interviews and medical records were utilized to assess diabetes (gestational and types 1 and 2), mode of delivery, parity, iron-deficiency anemia, infertility, endometriosis, maternal age, preterm delivery, and birthweight.^30^ Gestational age at delivery was estimated using an algorithm based on the last menstrual period and <20 weeks gestation ultrasound results.^34^

### Statistical Analysis

Participant characteristics were first compared descriptively by UF and HDP status. Continuous variables were assessed using two-sample t-tests, while categorical and binary variables were compared using Chi-squared tests. Plasma biomarker concentrations were compared between women with and without UFs and HDPs using Wilcoxon rank-sum tests. In all the models in this study, covariates controlled for included maternal age, race, BMI, parity, delivery type, diabetes, iron-deficiency anemia, and endometriosis. Complete-case analyses were used throughout, resulting in reduced sample sizes for analyses involving homocysteine due to missing laboratory data.

Vitamin D, vitamin B12, folate, and homocysteine were analyzed both continuously and categorically as quartiles. This was done due to the lack of consensus in the literature regarding cutoffs for vitamin sufficiency.^36,37^ Subsequently, for each plasma biomarker quartile, the odds of developing HDPs and UFs were calculated. Odds ratios (ORs) are reported with their respective 95% confidence intervals (CIs), and significance was determined by a 95% CI that did not contain 1. For folate, vitamin B12, and vitamin D, the higher-concentration group (Q4) served as the reference, whereas for homocysteine, the lowest-concentration group (Q1) served as the reference, reflecting our hypothesis that elevated homocysteine is associated with increased UF and HDP risk.

Pearson correlation coefficients (r) were calculated between homocysteine and vitamin B12, as well as homocysteine and folate, stratified by UF and HDP status.

To facilitate interaction and joint-effects analyses, quartile-level results were used to create dichotomized categories for each plasma biomarker. By default, plasma biomarkers were divided at the median to create balanced groups (“Low” or “High” vs. “Adequate”). For vitamin D, folate, and vitamin B12, lower concentrations were considered potentially adverse and were categorized as “Low” versus “Adequate” (reference). For homocysteine, higher concentrations were considered potentially adverse and were categorized as “Adequate” (reference) versus “High,” reflecting its hypothesized pathogenic role. When quartile-level analyses revealed statistically significant differences, cut-points were instead defined to reflect those contrasts (e.g., isolating the extreme quartile associated with higher or lower risk).

The potential modifying effect of each plasma biomarker on the association between UFs (no vs. yes) and HDPs (no vs. yes) was evaluated using binomial generalized linear models. We constructed one model for each plasma biomarker. The analytical model included HDP status as the dependent variable, UF status as the independent variable, dichotomized nutritional category of the plasma biomarker as an effect modifier, an interaction term between UFs and the biomarker, and the aforementioned covariates. The statistical significance of the interaction term was examined to determine whether plasma biomarker status modified the association between UFs and HDPs.

To further examine the combined influence of UFs and nutrient categories, composite exposure variables were created to represent all four combinations of UF status (Yes vs. No) and dichotomized plasma biomarker categories. These composite variables were entered as predictors in binomial generalized linear models with HDP as the outcome, adjusting for the same covariates. ORs and 95% CIs were estimated to compare the relative odds of HDPs across composite exposure groups.

The threshold for statistical significance for all tests was p<0.05. All statistical analysis was performed using R (version 4.5.1).

## Results

Seven hundred and twenty-two women with complete information regarding HDP status, UF status, cord vitamin D, plasma B12, and plasma folate were included in this analysis (Table 1). Of those, 12% had UFs, whereas 88% did not. 90% of women in this analysis did not have an HDP. 10% of women did have an HDP, of which 13.9% had gestational hypertension, 44.4% had PE without severe features, 40.3% had severe PE, and 5.6% had HELLP Syndrome; categories were not mutually exclusive.

**Table 1:** Study participant characteristics by uterine fibroid and hypertensive disorder of pregnancy status. Values are presented as mean (SD) or n(%). Due to rounding, not all percentages sum to 100%. Some patients had >1 HDP diagnosis.

| Demographic | Uterine Fibroids (UFs) |  |  | Hypertensive Disorders of Pregnancy (HDPs) |  |  |
| --- | --- | --- | --- | --- | --- | --- |
|  | No (n=635) | Yes (n=87) | p-value | No (n=650) | Yes (n=72) | p-value |
| Maternal Age | 28 (6) | 32 (6) | **<0.001** | 28 (7) | 31 (6) | **0.009** |
| Race/Ethnicity |  |  | **<0.001** |  |  | 0.117 |
| Black/African American | 380 (60%) | 71 (82%) |  | 406 (62%) | 45 (63%) |  |
| Hispanic | 151 (24%) | 9 (10%) |  | 147 (23%) | 13 (18%) |  |
| White | 49 (7.7%) | 1 (1.1%) |  | 47 (7.2%) | 3 (4.2%) |  |
| Other | 55 (8.7%) | 6 (6.9%) |  | 50 (7.7%) | 11 (15%) |  |
| BMI | 26.1 (6.1) | 28.6 (6.6) | **0.002** | 26.1 (6.1) | 29.1 (5.9) | **<0.001** |
| Unknown | 27 | 6 |  | 28 | 5 |  |
| Educational Level |  |  | 0.892 |  |  | 0.861 |
| Any College | 222 (35%) | 29 (34%) |  | 227 (35%) | 24 (33%) |  |
| No College | 410 (65%) | 57 (66%) |  | 419 (65%) | 48 (67%) |  |
| Unknown | 3 | 1 |  | 4 | 0 |  |
| Parity |  |  | 0.190 |  |  | 0.562 |
| Non-Nulliparous | 380 (60%) | 59 (68%) |  | 398 (61%) | 41 (57%) |  |
| Nulliparous | 255 (40%) | 28 (32%) |  | 252 (39%) | 31 (43%) |  |
| Maternal Smoking at any Point During Pregnancy | 114 (18%) | 13 (15%) | 0.580 | 113 (18%) | 14 (20%) | 0.739 |
| Unknown | 9 | 1 |  | 8 | 2 |  |
| Gestational Age at Delivery (weeks) | 38.61 (2.48) | 37.94 (3.18) | 0.060 | 38.71 (2.39) | 36.90 (3.50) | **<0.001** |
| Preterm Delivery (<37 weeks) | 107 (17%) | 19 (22%) | 0.318 | 95 (15%) | 31 (43%) | **<0.001** |
| Birthweight (grams) | 3,115 (633) | 3,128 (772) | 0.876 | 3,154 (610) | 2,781 (881) | **<0.001** |
| Mode of Delivery |  |  | **<0.001** |  |  | **0.034** |
| Cesarean Section | 180 (28%) | 46 (53%) |  | 195 (30%) | 31 (43%) |  |
| Vaginal Delivery | 453 (72%) | 41 (47%) |  | 453 (70%) | 41 (57%) |  |
| Unknown | 2 | 0 |  | 2 | 0 |  |
| Diabetes Status |  |  | 0.071 |  |  | **<0.001** |
| Gestational Diabetes | 26 (4.1%) | 6 (6.9%) |  | 24 (3.7%) | 8 (11%) |  |
| Preconception Diabetes | 14 (2.2%) | 5 (5.7%) |  | 13 (2.0%) | 6 (8.3%) |  |
| None | 593 (94%) | 76 (87%) |  | 611 (94%) | 58 (81%) |  |
| Unknown | 2 | 0 |  | 2 | 0 |  |
| Iron-Deficiency Anemia | 49 (8.2%) | 25 (29%) | **<0.001** | 66 (11%) | 8 (12%) | 0.924 |
| Unknown | 40 | 0 |  | 35 | 5 |  |
| Endometriosis | 0 (0%) | 4 (4.6%) | **<0.001** | 4 (0.7%) | 0 (0%) | >0.999 |
| Unknown | 40 | 0 |  | 35 | 5 |  |

Values represented as Mean (SD) or n (%)

Women with UFs were significantly older (32 vs. 28 years) and had significantly higher BMI (28.6 vs. 26.1 kg/m^2^) compared with women without UFs; similarly, women with HDPs were significantly older (31 vs. 28 years) and had significantly higher BMI (29.1 vs. 26.1 kg/m^2^) than those without HDPs. Race and ethnicity distributions differed by UF status, with a higher proportion of Black/African American women among those with UFs (82% vs. 60%), but did not differ by HDP status. Women with HDPs delivered at significantly earlier gestational ages (36.9 vs. 38.7 weeks), had significantly lower infant birthweights (2,781 vs. 3,154 g), higher rates of preterm delivery (43% vs. 15%), and were more likely to undergo cesarean sections (43% vs. 30%). Cesarean-sections were also significantly more common among women with UFs (53% vs. 28%). Iron-deficiency anemia and endometriosis were significantly more prevalent among women with UFs (29% vs. 8.2%, and 4.6% vs. 0%, respectively), whereas educational attainment, parity, and smoking during pregnancy did not differ by UF or HDP status.

### Distribution of Plasma Biomarkers

Distributions for vitamin D, vitamin B12, folate, and homocysteine are right-skewed (Figure 1). Median (IQR) concentrations of the analyzed plasma biomarkers were 12.90 (8.33-17.88) ng/mL for vitamin D, 260.50 (210.30-311.02) pmol/L for vitamin B12, 30.59 (20.49-47.40) nmol/L for folate, and 7.27 (5.98-9.25) μmol/L for homocysteine in the population overall. Amongst women with UFs and women with HDPs, there was no correlation between vitamin B12 and homocysteine. However, a weak positive correlation was identified for women without UFs (r=0.176, p<0.001, n=622) and women without HDPs (r=0.180, p<0.001, n=638), such that increased vitamin B12 levels were associated with increased homocysteine levels. There was no correlation between folate and homocysteine in any of the groups. Sample sizes were reduced in these calculations, as homocysteine data were unavailable for some participants.

**Figure 1:**
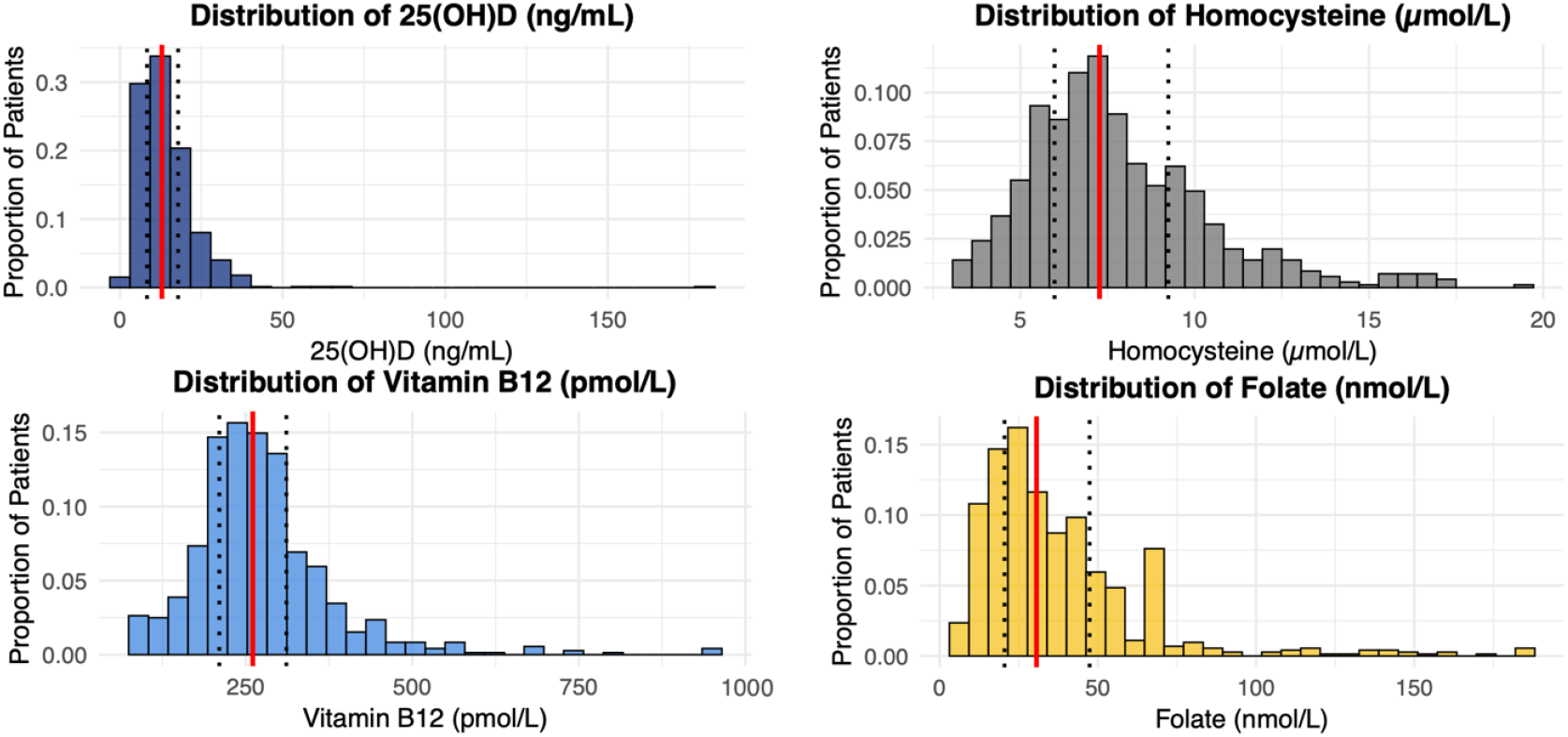
Distribution of plasma biomarker concentrations in postpartum (Day 1-2) maternal samples. Solid red lines indicate sample medians, and dotted lines denote quartiles 1 and 3.

### Plasma Biomarkers and the Relationship to UFs and HDPs

Mean (SD) plasma biomarker concentrations in women with and without uterine fibroids and hypertensive disorders of pregnancy are presented in Table 2. Concentrations of vitamin D, vitamin B12, folate, and homocysteine did not differ significantly between women with and without UFs after covariate adjustment (p>0.05 for all). In comparison, women with HDPs had significantly higher homocysteine concentrations than those without HDPs (p=0.028). This was also true when analyzed by quartile, with no significant differences in odds of UFs observed across quartiles of vitamin D, vitamin B12, folate, or homocysteine (Table 3).

**Table 2:** Plasma biomarker concentrations in women with vs. without uterine fibroids and hypertensive disorders of pregnancy. Values represent unadjusted mean (SD).

| Plasma Biomarker | Uterine Fibroids (UFs) |  |  | Hypertensive Disorders of Pregnancy (HDPs) |  |  |
| --- | --- | --- | --- | --- | --- | --- |
|  | No (n=635) | Yes (n=87) | p-value | No (n=650) | Yes (n=72) | p-value |
| *Cord blood* |  |  |  |  |  |  |
| 25(OH)D (ng/mL) | 14.5(10.7) | 13.9(7.0) | 0.869 | 14.5(10.6) | 13.9(7.2) | 0.829 |
| *Maternal blood* |  |  |  |  |  |  |
| Vitamin B12 (pmol/L) | 274.0 (110.0) | 277.0(94.8) | 0.339 | 273.0(110.0) | 281.0(92.1) | 0.177 |
| Folate (nmol/L) | 38.6(28.3) | 34.0(19.4) | 0.295 | 38.5(27.5) | 33.9(26.0) | 0.066 |
| Homocysteine (μmol/L) | 7.9(2.7) | 7.4(2.3) | 0.196 | 7.7(2.5) | 8.6(3.3) | **0.028** |

Values represent the mean(SD)

**Table 3:** Odds ratios for developing uterine fibroids or hypertensive disorders of pregnancy by plasma biomarker quartile.

| Plasma Biomarker | Quartile Number | Quartile Range (Min, Max) | Uterine Fibroids (UFs) | Hypertensive Disorders of Pregnancy (HDP) |
| --- | --- | --- | --- | --- |
|  |  |  | Odds Ratio (95% Confidence Interval) | Odds Ratio (95% Confidence Interval) |
| 25(OH)D (ng/mL) | 1 | (1.3, 8.3) | 0.81(0.41,1.60) | 0.88(0.42, 1.83) |
|  | 2 | (8.4, 12.9) | 0.95(0.49,1.83) | 0.72(0.33,1.55) |
|  | 3 | (12.9, 17.9) | 0.81(0.41,1.61) | 0.94(0.46, 1.94) |
|  | 4 | (18.0, 181.1) | Reference | Reference |
| Vitamin B12 (pmol/L) | 1 | (99.4, 210.1) | 0.63(0.31,1.23) | 0.57(0.25,1.27) |
|  | 2 | (210.8, 260.9) | 0.71(0.36,1.37) | 0.94(0.46,1.93) |
|  | 3 | (261.0, 311.2) | 0.79(0.41,1.51) | 1.06(0.52,2.14) |
|  | 4 | (311.2, 960.2) | Reference | Reference |
| Folate (nmol/L) | 1 | (6.6, 20.5) | 1.64(0.84,3.30) | **2.35(1.13,5.12)** |
|  | 2 | (20.5, 30.6) | 1.18(0.58,2.45) | 1.08(0.45,2.57) |
|  | 3 | (30.7, 47.4) | 1.12(0.54,2.35) | 1.34(0.59,3.10) |
|  | 4 | (47.5, 185.5) | Reference | Reference |
| Homocysteine (μmol/L) | 1 | (3.1,6.0) | Reference | Reference |
|  | 2 | (6.0, 7.3) | 1.15(0.60, 2.22) | 1.40(0.58, 3.47) |
|  | 3 | (7.3, 9.2) | 0.76(0.37, 1.53) | 1.92(0.85, 4.59) |
|  | 4 | (9.3, 19.3) | 0.81(0.40, 1.63) | **2.36(1.07, 5.53)** |

In contrast, differences by quartile were noted for HDPs; compared with women in the highest quartile of folate, those in the lowest quartile (Q1) had higher odds of HDPs (adjusted OR 2.35, 95% CI 1.13–5.12). Similarly, women in the highest quartile (Q4) of homocysteine had higher odds of HDPs (adjusted OR 2.36, 95% CI 1.07–5.53) compared with those in the lower quartiles. Taken together, these analyses indicate low folate and elevated homocysteine levels were independently associated with higher odds of HDPs. These findings suggest a threshold-based relationship for folate and homocysteine, motivating subsequent dichotomized analyses. Folate was categorized as “Low” (Q1) versus “Adequate” (Q2–Q4) and homocysteine as “Adequate” (Q1–Q3) versus “High” (Q4). No associations were found between HDPs and either vitamins D or B12.

### Interaction and joint-effects analyses of UFs, plasma biomarkers, and HDPs

No statistically significant interaction effects were observed between UFs and any plasma biomarker concentration in relation to HDPs (Supplemental Table 1). Joint-effects models were also used to assess the combined influence of UFs and plasma biomarkers on the odds of HDPs (Table 4). Across most plasma biomarkers, no significant joint associations were detected. However, women with UFs and low folate concentrations had significantly higher odds of HDPs compared to those without UFs and with adequate folate (adjusted OR=1.84, 95% CI 1.07–3.17, p=0.027). No significant joint effects were observed for vitamin D, vitamin B12, or homocysteine.

**Table 4:**
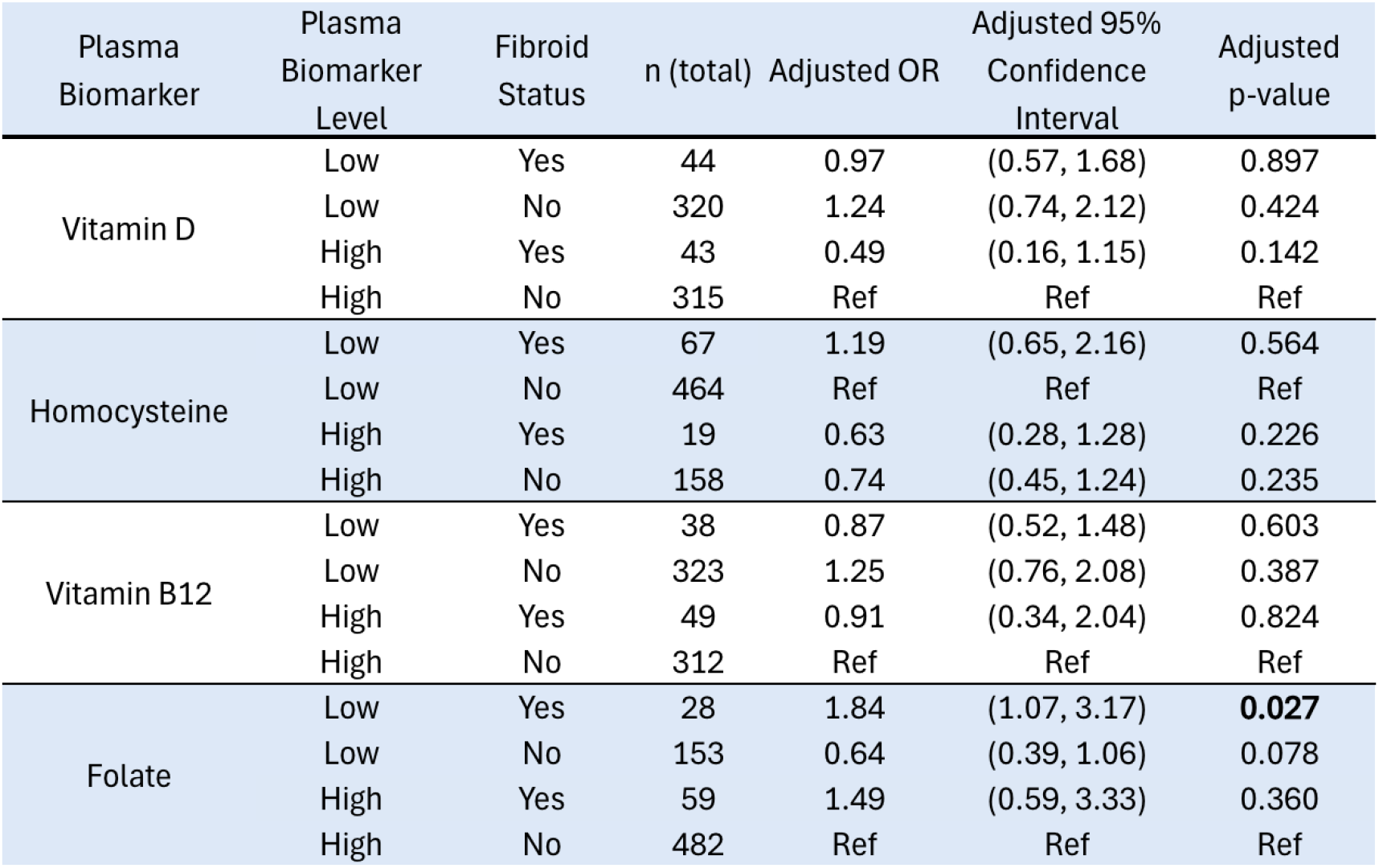
Joint effects of uterine fibroid status and plasma biomarker levels on the odds of hypertensive disorders of pregnancy.

| Plasma Biomarker | Plasma Biomarker Level | Fibroid Status | n (total) | Adjusted OR | Adjusted 95% Confidence Interval | Adjusted p-value |
| --- | --- | --- | --- | --- | --- | --- |
| Vitamin D | Low | Yes | 44 | 0.97 | (0.57, 1.68) | 0.897 |
|  | Low | No | 320 | 1.24 | (0.74, 2.12) | 0.424 |
|  | High | Yes | 43 | 0.49 | (0.16, 1.15) | 0.142 |
|  | High | No | 315 | Ref | Ref | Ref |
| Homocysteine | Low | Yes | 67 | 1.19 | (0.65, 2.16) | 0.564 |
|  | Low | No | 464 | Ref | Ref | Ref |
|  | High | Yes | 19 | 0.63 | (0.28, 1.28) | 0.226 |
|  | High | No | 158 | 0.74 | (0.45, 1.24) | 0.235 |
| Vitamin B12 | Low | Yes | 38 | 0.87 | (0.52, 1.48) | 0.603 |
|  | Low | No | 323 | 1.25 | (0.76, 2.08) | 0.387 |
|  | High | Yes | 49 | 0.91 | (0.34, 2.04) | 0.824 |
|  | High | No | 312 | Ref | Ref | Ref |
| Folate | Low | Yes | 28 | 1.84 | (1.07, 3.17) | **0.027** |
|  | Low | No | 153 | 0.64 | (0.39, 1.06) | 0.078 |
|  | High | Yes | 59 | 1.49 | (0.59, 3.33) | 0.360 |
|  | High | No | 482 | Ref | Ref | Ref |

## Discussion

Women with UFs face a greater risk for HDPs, which are linked to numerous adverse maternal outcomes, including hepatic dysfunction, elevated stroke risk, and acute renal failure.^1,7^ The effects of HDPs may persist beyond the postpartum period, leading to an increased risk of premature maternal mortality, particularly due to cardiovascular disease.^8^ Micronutrient supplementation is theoretically a possible intervention to attenuate the risk women with UFs face for developing HDPs, as they may help address the shared pathophysiology underlying HDPs and UFs.

Our study evaluated the independent and combined effects of micronutrient status and fibroid presence, including potential effect modification, on HDP risk and found that neither micronutrients nor homocysteine levels modified the association between UFs and HDPs. However, the joint-effects models identified higher odds of HDPs among women with both fibroids and low folate compared with those with either exposure alone. These findings highlight the need to further investigate folate’s potential role as a modifiable factor that is associated with lower odds for HDPs among women with UFs.

In our cohort, increased maternal age and BMI were associated with UFs and HDPs. Gestational/preconception diabetes was associated with women with HDPs. These covariates reflect known risk factors for HDPs and UFs.^42–45^ Iron-deficiency anemia and endometriosis were found to be more prevalent in women with UFs, aligning with previous studies describing the comorbidity between UFs and both endometriosis and iron-deficiency anemia.^46,47^ The consistency between our cohort’s characteristics and known UF and HDP associations supports the validity of the studied population.

Consistent with prior studies, there was no association between UFs and folate or vitamin B12 levels. However, in contrast to previous reports, we did not find an association between lower vitamin D quartiles and UF risk. One possible explanation is that our cohort predominantly consists of Black and Hispanic women, who are more likely to be vitamin D deficient compared to non-Hispanic White women, potentially limiting variability in vitamin D exposure.^48,49^ Similarly, Baird et al. found that only 10% of Black women had sufficient vitamin D concentrations (defined as >20ng/ml) compared to 50% of White women.^50^ However, Baird et al. still found that vitamin D sufficiency was associated with lower odds of UFs in the overall cohort (adjusted OR 0.68, 95% CI 0.48–0.96), with race-stratified analyses demonstrating similar inverse associations between circulating vitamin D concentrations and UF risk among Black and White women. This suggests that there are additional unmeasured variables, such as level of sun exposure, that may confound the relationship between vitamin D and UFs in our study.

Homocysteine induces oxidative stress that damages endothelial cells and contributes to the pathophysiology of HDPs. Previous studies have reported elevated homocysteine associated with HDPs, albeit with mixed findings regarding the association between folate and HDPs.^51–53^ Folate functions as an essential cofactor in homocysteine metabolism; thus, insufficient folate impairs this pathway and leads to homocysteine accumulation. In this study, we observed that patients in the lowest quartile of folate concentrations had higher odds of developing an HDP compared to women in the highest folate quartile (adjusted OR 2.35, 95% CI 1.13-5.12). Similarly, we found women in the highest quartile of homocysteine had higher odds of HDPs (adjusted OR 2.36, 95% CI 1.07-5.53). These findings are biologically concordant, as both low folate and elevated homocysteine reflect increased oxidative stress, which can contribute to HDP development.

Although low folate concentrations are biologically expected to correlate with higher homocysteine levels, thereby increasing HDP risk, no correlation was observed in our cohort. Folate and homocysteine levels change during pregnancy, potentially driven by endocrine changes, folic acid supplementation, hemodilution, renal clearance alterations, and decreases in serum albumin.^54,55^ These adaptations during pregnancy may have obscured the anticipated inverse association between folate and homocysteine.

The strengths of this study include its use of both continuous and categorical analyses, enabling assessment of plasma biomarker relationships while reducing the influence of outliers that may skew results. The cohort composition, represented predominantly by Black and Hispanic women, provides data that is generalizable to the women facing the greatest morbidity and mortality due to HDPs. While this is inherently a strength, the results are less generalizable to White populations.

This study has various limitations. Because a retrospective analysis was performed, we had to exclude many patients in the cohort who did not have complete vitamin, UF, and/or HDP data. As a result, the sample sizes for the cases of UFs and HDPs were diminished, resulting in decreased statistical power. This study was also an observational, rather than an experimental study. Although HDPs encompass a broader spectrum of diseases ranging from gestational hypertension to HELLP syndrome, 84.7% of our patients had PE with or without severe features. This limits the generalizability of our results to disorders beyond PE, as we did not have sufficient sample sizes to study the associations between micronutrients and HDP subtypes.

Another limitation is the use of a single measurement of plasma biomarkers, which may vary over the course of pregnancy due to physiologic and metabolic changes. Biomarker levels were therefore assumed to be stable over time, which may have influenced estimated associations. Although UF and HDP status were established prior to biomarker assessment, maternal and cord blood vitamin concentrations were assumed to reflect overall pregnancy micronutrient status.

Altogether, women with UFs face heightened risk for HDPs, which carry lasting maternal health consequences. Although micronutrients did not modify this association, the observed joint effects of low folate and UFs highlight a potentially modifiable risk factor that warrants further investigation. Experimental studies are needed to determine whether targeted nutritional interventions can reduce HDP-related morbidity, particularly in populations bearing the greatest burden of disease.

## Data Availability

Requests for access to de-identified individual participant-level data referred to in this manuscript will be considered by the authors, subject to institutional policies, IRB approval, informed consent limitations, and completion of any required data use agreements.

## Funding

The Boston Birth Cohort (the parent study) is supported in part by the National Institutes of Health (NIH) grants (2R01HD041702, R01HD098232, R21AI154233, R01ES031272, R01ES031521, and U01 ES034983) and by the Health Resources and Services Administration (HRSA) of the U.S. Department of Health and Human Services (HHS) cooperative agreement (UT7MC45949). This information or content and conclusions are those of the authors and should not be construed as the official position or policy of, nor should any endorsements be inferred by the funding agencies.

## Role of the Funder/Sponsor

The funding organizations had no role in the design and conduct of the study; collection, management, analysis, and interpretation of the data; preparation, review, or approval of the manuscript; and decision to submit the manuscript for publication.

## Conflicts of Interests

The authors report no conflicts of interest.

## Acknowledgements

We gratefully acknowledge Serena Rusk, MPH, BS and Xiumei Hong, MD, PhD for their support throughout the development of this study.

## Supplemental Data

**Table 1:** Interaction between uterine fibroid status and plasma biomarker levels on the odds of hypertensive disorders of pregnancy.

| Plasma Biomarker | Adjusted Odds Ratio for Interaction Term (95% Confidence Interval) | Adjusted p-value |
| --- | --- | --- |
| 25(OH)D | 0.77(0.50, 1.14) | 0.212 |
| Vitamin B12 | 0.94 (0.64, 1.41) | 0.760 |
| Folate | 1.03 (0.69, 1.52) | 0.899 |
| Homocysteine | 1.15 (0.76, 1.73) | 0.491 |

## Notes

### Competing Interest Statement

The authors have declared no competing interest.

### Author Declarations

IRB of Johns Hopkins School of Public Health gave ethical approval for this work

